# The influence of serum Luteinising hormone concentrations post GnRH agonist trigger on metaphase II oocytes in IVF/ICSI cycles

**DOI:** 10.1101/2025.05.28.25328348

**Authors:** Shital Sawant, Sasha Tulsyan, Roy Homburg

## Abstract

**Background:** Gonadotropin releasing hormone agonist (GnRHa) trigger is increasingly used in IVF/ICSI cycles to mitigate the risk of ovarian hyperstimulation syndrome (OHSS), particularly in women with polycystic ovary syndrome (PCOS). While safer than hCG, the endocrine response to GnRHa is highly variable. It has been hypothesised that there exists a threshold serum luteinizing hormone (LH) concentration post trigger that is predictive of optimal oocyte maturation.

However, this threshold has not been clearly established. This study aimed to identify optimum and minimum LH levels measured 12 hours after GnRHa trigger that are predictive of mature oocyte (metaphase II, MII) yield in PCOS patients.

**Methods:** This prospective, single centre observational study included 104 women with PCOS undergoing controlled ovarian stimulation using a GnRH antagonist protocol. Final oocyte maturation was induced with 2mL Buserelin. Serum luteinising hormone, follicle stimulating hormone (FSH), and progesterone were measured 12 hours post trigger. The primary outcome was correlation between hormone levels and MII oocyte yield. Secondary outcomes included fertilisation and blastocyst conversion. Pearson correlation analysis and receiver operating characteristic (ROC) curves were used to assess the predictive power of hormone levels for oocyte maturity.

**Results:** The mean LH level at 12 hours post trigger was 34.05 (SD 23.07 IU/L, range: 0.9 to 134.4 IU/L). No statistically significant correlation was found between LH and MII oocyte yield (r = negative 0.123; p = 0.215), fertilisation rate (r = 0.046; p = 0.641), or blastocyst development (r = 0.162; p = 0.101). Similarly, FSH and progesterone showed no significant correlations with primary outcomes. ROC analysis revealed poor predictive capacity of post trigger LH for mature oocyte yield (AUC = 0.402). Notably, patients with 100% MII yield had LH levels as low as 6.8 IU/L, while those with poor MII yield (>0 to 25%) had LH levels as high as 78.9 IU/L, indicating no clear minimum or optimum LH threshold for predicting mature oocyte development.

**Conclusion:** No specific LH thresholds at 12 hours post GnRHa trigger could be identified as predictive of optimal or suboptimal oocyte maturation. Routine post trigger LH hormone level measurement lacks clinical utility in predicting mature oocyte yield in PCOS IVF/ICSI cycles.

## INTRODUCTION

Human chorionic gonadotrophin (hCG) is structurally and functionally similar to LH. This feature means that it has long been used as a surrogate trigger for the LH peak to achieve final oocyte maturation in IVF cycles. However, the administration of hCG causes a sustained luteotrophic effect (1) since it has a greater half-life (>24hours) than LH (<60minutes). This prolonged luteotrophic activity stimulates the production of vascular endothelial growth factor (VEGF) thereby significantly increasing intravascular permeability and the risk of developing ovarian hyperstimulation syndrome (OHSS) (2,3).

OHSS remains an important iatrogenic complication of controlled ovarian stimulation in IVF. The incidence varies from 33% in mild to 2-8 % in moderate to severe forms (4). Polycystic ovary syndrome (PCOS) is a significant risk factor for the development of OHSS with one systematic review suggesting an odds ratio as high as 6.8 (5). It is of further significance as PCOS is a common condition with a prevalence of 5-18% (6) and can cause anoluvation with sub or infertility. As such it makes up a key demographic amongst women seeking fertility treatments.

In aims to reduce the risk of OHSS during IVF cycles, GnRH agonist (GnRHa) trigger can be used as an alternative to hCG. The effect of GnRHa more closely mimics the endogenous LH and FSH surges seen in natural cycles, has shorter lasting effects, and causes considerably lower levels of VEGF production (7). Evidence from multiple trials spanning the last decade suggests that using GnRHa as an ovulation trigger significantly reduces, or even eliminates, the incidence of OHSS (8–11).

There is however a concern that the use of GnRHa rather than hCG can result in lower mature oocyte yield. The LH surge induced by GnRHa consists of two phases – a short ascending limb (>4hours) and a long descending limb (>20hours), lasting 24-36 hours on average (12). On the other hand, the natural mid-cycle surge of LH has three phases – a rapidly ascending phase (14hours), a plateau phase (14 hours) and a descending phase (20hours), lasting 48 hours (13). Thus, compared to a natural cycle, the total amount and duration of gonadotrophin action is reduced when using GnRHa trigger.

The LH surge post GnRHa trigger is essential in stimulating oocyte maturity prior to egg retrieval in IVF cycles. Thus, it is possible that a suboptimal pituitary LH response to GnRHa occurs in some patients, potentially reducing mature oocyte yield and increasing the risk of empty follicle syndrome (EFS). There may be a threshold of post-trigger LH level, above which a maximal oocyte cohort is typically obtained, and below which oocyte yield and maturity tend to decline. There may also be a threshold of clinical relevance to cycle outcome, below which dramatic declines in yield and maturity may occur (14).

Shapiro et al. demonstrated that an LH level of <52 IU/l at 12 hours post GnRHa trigger was suboptimal while a LH level of <12 IU/l was inadequate for oocyte yield and maturity (15). In the same study, a LH level of <12.0 IU/l provided a 50% sensitivity and 97% specificity in predicting low yield and achieved 38% sensitivity and 97% specificity in predicting low oocyte maturity. Similarly, Chen et al. showed that at LH levels <15.0 IU/l there was a statistically significant reduction in oocyte yield, although oocyte maturation and fertilization rates were not affected (16).

In this study, we measured serum LH, FSH, and progesterone levels 12 hours post GnRH agonist trigger during standard antagonist protocol IVF/ICSI cycles. The aim was to identify optimum levels at which matured (metaphase II, MII) oocytes are obtained and the levels below which, the possibility of obtaining MII oocytes is small or negligible. The study would also help determine if there is a required minimum serum concentration of these hormones to define optimal outcomes for oocyte maturity and/or pregnancy outcome. These threshold levels would in turn help decide the ideal dosage of GnRH agonist trigger for a given patient to reach a required post-trigger threshold and attain the optimal outcome. Whether these dosages correspond or change with age, AMH, AFC, or BMI would be further areas of research.

## MATERIALS AND METHODS

### Study Population and Design

This was a single centre prospective observational study at a university fertility center in northeast London, UK. The study recruited 104 women undergoing IVF/ICSI treatment from April 2019 to April 2021 . All patients had a diagnosis of PCOS with AMH >20 pmol/L or AFC >/= 20, and received a GnRH agonist as the trigger for oocyte maturation. All patients were pre-assessed by a fertility specialist and screened for study suitability. This study was interrupted by the SARS-CoV-2 pandemic and as a result, the number of study participants were less than originally intended.

### Patients ’consent

This study received approval through Integrated Research Application System (IRAS) in the NHS UK. All patients received written information regarding the study and OHSS prior to beginning. All included patients gave informed, written consent to be part of the study.

### Stimulation and Trigger Protocol

All patients had their first ultrasound scan between first and fourth day of their menstrual cycle. The GnRH antagonist protocol was used, with FSH dose based on AMH level – either 125, 225 or 300 IU (pre-decided at pre-assessment appointment). Controlled ovarian hyperstimulation (COH) was carried out until the leading follicle had reached >17mm in size. GnRH agonist Suprecur(2ml Buserelin) trigger was given 35-36 hours prior to planned egg collection timing. Blood samples for LH, FSH and progesterone levels were collected at time of trigger and 12 hours post trigger.

Transvaginal ultrasonography was utilised to guide ovum pickup performed 34–36 hours after GnRH agonist administration. Depending on the quality of sperm, oocytes were inseminated via conventional IVF or ICSI. Embryos were routinely monitored in real time to confirm successful fertilisation and susbsequently assessed and graded according to Gardner and Schoolcraft Grading system. Cryopreserved embryos were then stored for use in a subsequent frozen embryo transfer (FET) cycle at a later stage.

The primary end points were an optimal level of LH 12h post trigger which produces the highest proportion of metaphase II oocytes collected and the level of LH below which no oocytes were collected. We reviewed and correlated the levels of progesterone and FSH with number of MII oocytes.

### Hormone Measurement

An electrochemiluminescence immunoassay (ECLIA) technology, utilizing the Elecsys Hormone Measurement System by Roche, was employed to assess hormone levels. This system was used to quantify Luteinizing Hormone (LH), Follicle-Stimulating Hormone (FSH), Estradiol (E2), and Progesterone (P4) levels. AMH assays were performed using the Beckman-Coulter Generation II assay and values presented in pmol/L.

### Statistical Analysis

LH, FSH, and progesterone levels were measured at 12 hours post GnRH agonist administration and compared to outcomes for percentage of mature oocytes, fertilisation rate, and blastocyst formation rate. The results were analysed using SPSS software and Pearson correlation to determine the relationship between hormone levels and outcome.

## RESULTS

A total of 104 patients were included in the study and results. The mean age of the cohort was 33.5 years (range of 24-43 years) with a mean BMI of 24.9kg/m^2^ (range of 18.5-34.2kg/m^2^). 65 patients underwent IVF and 38 patients underwent ICSI. One patient failed to respond to controlled ovarian hyperstimulation and therefore did not have IVF or ICSI as there were no eggs present.

The mean LH level 12 hours post trigger was 34.05IU/L (SD+/-23.07IU/L). There was a wide range of levels with the lowest being 0.9IU/L and highest being 134.4IU/L. The total number of mature eggs collected for each patient was recorded. The mature oocyte yield was then calculated as a percentage of mature oocytes within the total number of eggs collected for each patient. Out of the 103 patients that responded to COH, 4 showed a mature oocyte yield 0-25%, 9 showed a yield of 25-50%, 28 showed a yield of 50-75% and 62 showed a high yield of 75-100%. Interestingly, the LH levels did not correspond with the number or percentage of mature oocytes and this data is represented in Figure 1.

**Figure 1.**
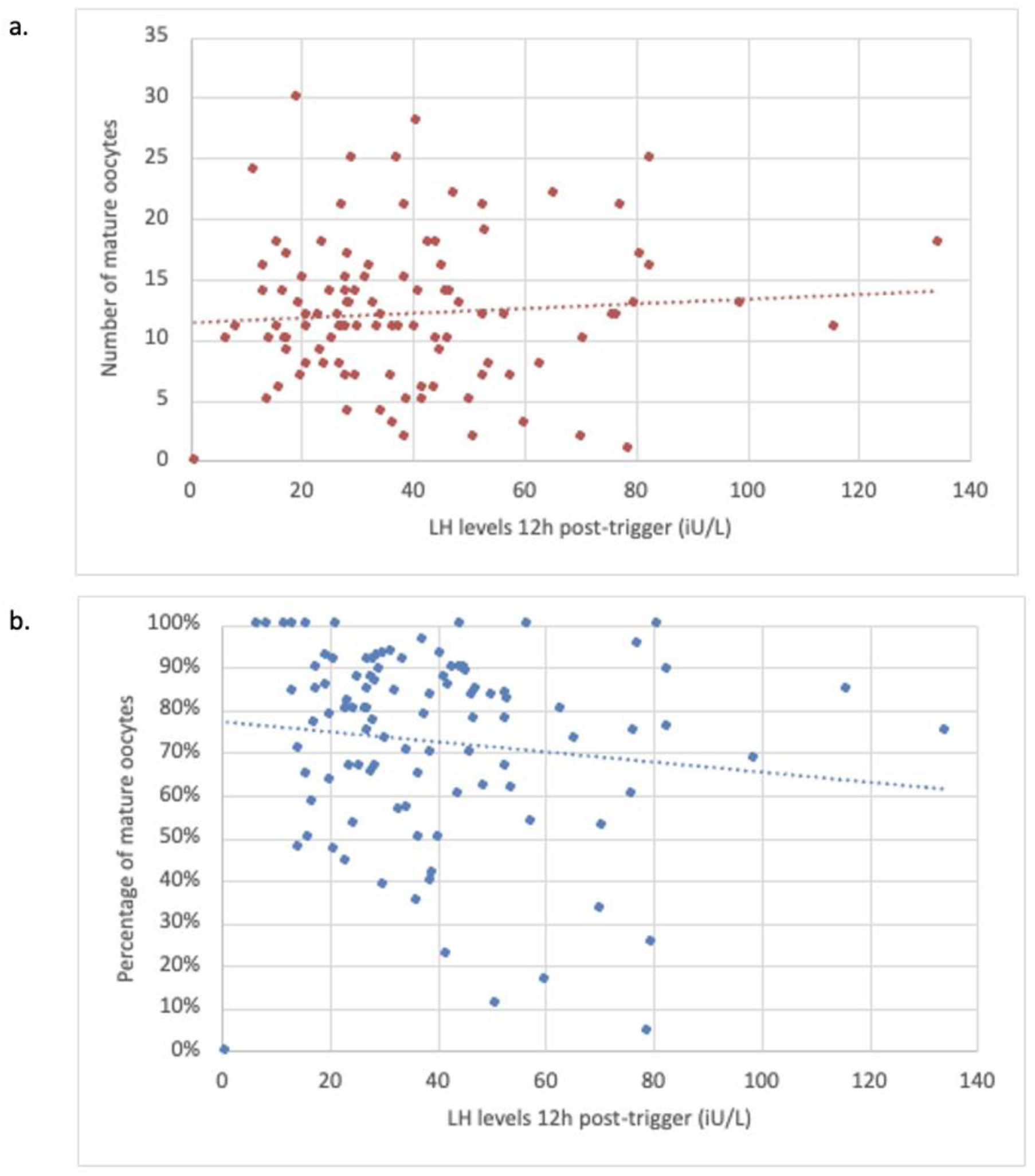
(a) Scatter plot showing LH level 12h post-trigger and the corresponding number of mature oocytes for that patient. (b) Scatter plot showing LH level 12h post-trigger and the corresponding percentage of mature oocytes for that patient.

SPSS statistical analysis software and Pearson correlation were then used to analyse the relationship between LH, FSH and progesterone levels and mature oocyte, fertilisation, and blastocyst rates. There was a minimal negative correlation between LH levels and MII oocytes (*r*=-0.123, *n*=104) however, this was not statistically significant (*p*=0.215). There was a minimal positive correlation between LH and fertilization and blastocyst development, respectively (*r*=0.046, *n*=104);*(r*=0.162, *n*=104) however, this was not statistically significant (*p*=0.641 and *p*=0.101, respectively). These results are shown in Table 1.

**Table 1.**
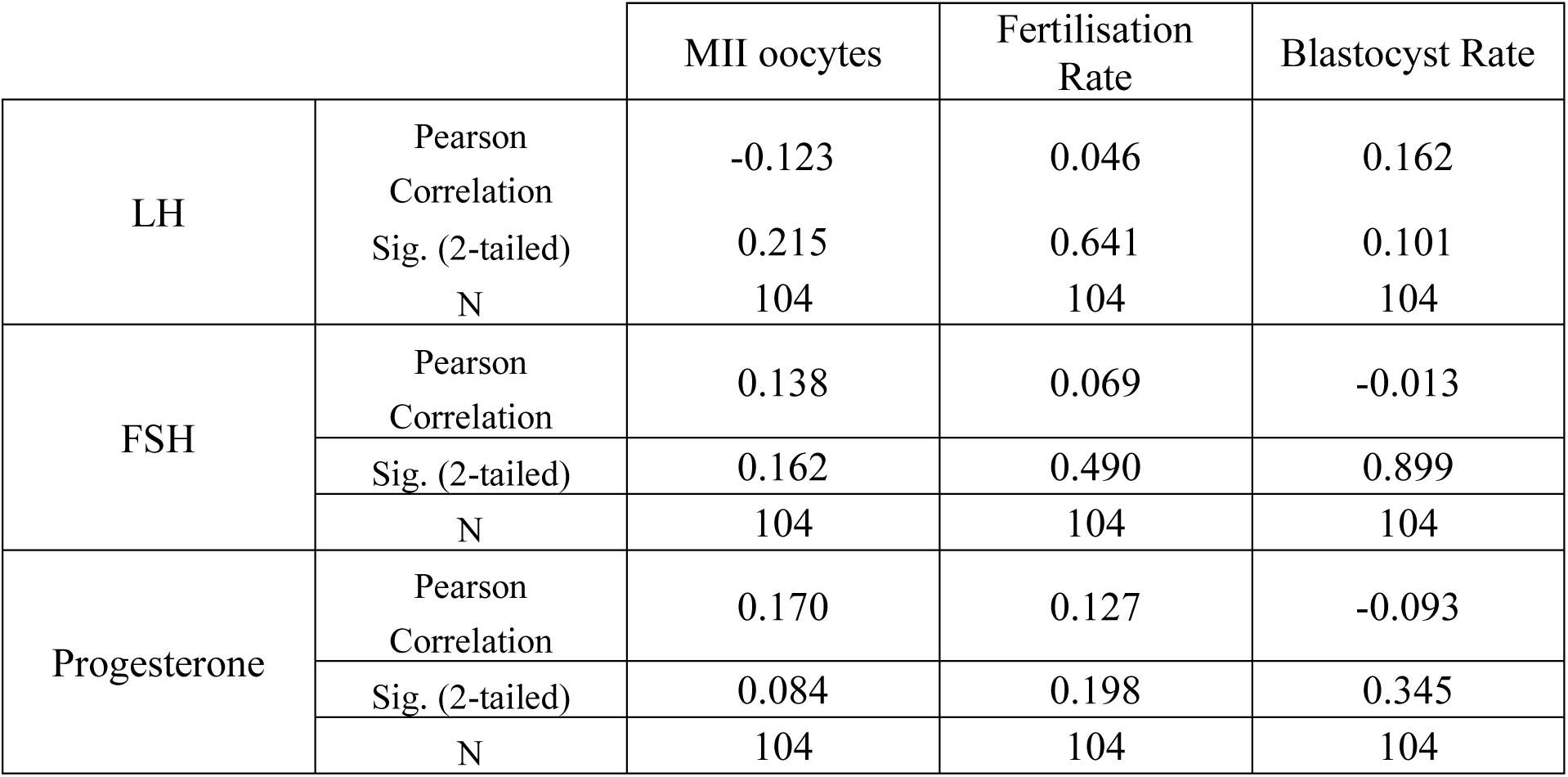
Pearson correlation between hormone levels and MII oocytes, fertilisation rate, and blastocyst rate.

There was a minimal positive correlation between FSH and MII oocytes and fertilization rate respectively (*r*=0.138, *n*=104);*(r*=0.069 *n*=104) however, this was not statistically significant (*p*=0.162; *p*=0.490). There was a minimal negative correlation between FSH and blastocyst development (*r*=-0.013, *n*=104) however, this was not statistically significant (*p*=0.899). These results are shown in Table 1.

There was a positive correlation between progesterone (P2) and MII oocytes and fertilization rate respectively (*r*=0.170, *n*=104);*(r*=0.127 *n*=104) however, this was not statistically significant (*p*=0.084; *p*=0.198). There was a negative correlation between P2 and blastocyst development (*r*=-0.093, *n*=104) however, this was not statistically significant (*p*=0.345). These results are shown in Table 1.

ROC curve analysis was performed to investigate the predictive performance of LH levels, 12hrs post trigger, for percentage MII oocyte yield, fertilization rate and blastocyst conversion rate. A statistically non-significant and limited predictive power was observed between percentage MII oocyte yield, fertilization rate and blastocyst conversion rate and LH levels 12hrs post trigger – AUC=0.402, 95% CI 0.27-0.53; AUC=0.573, 95% CI 0.43-0.71 and AUC=0.598, 95% CI 0.47-0.73 respectively. These results are shown in Figure 2.

**Figure 2.**
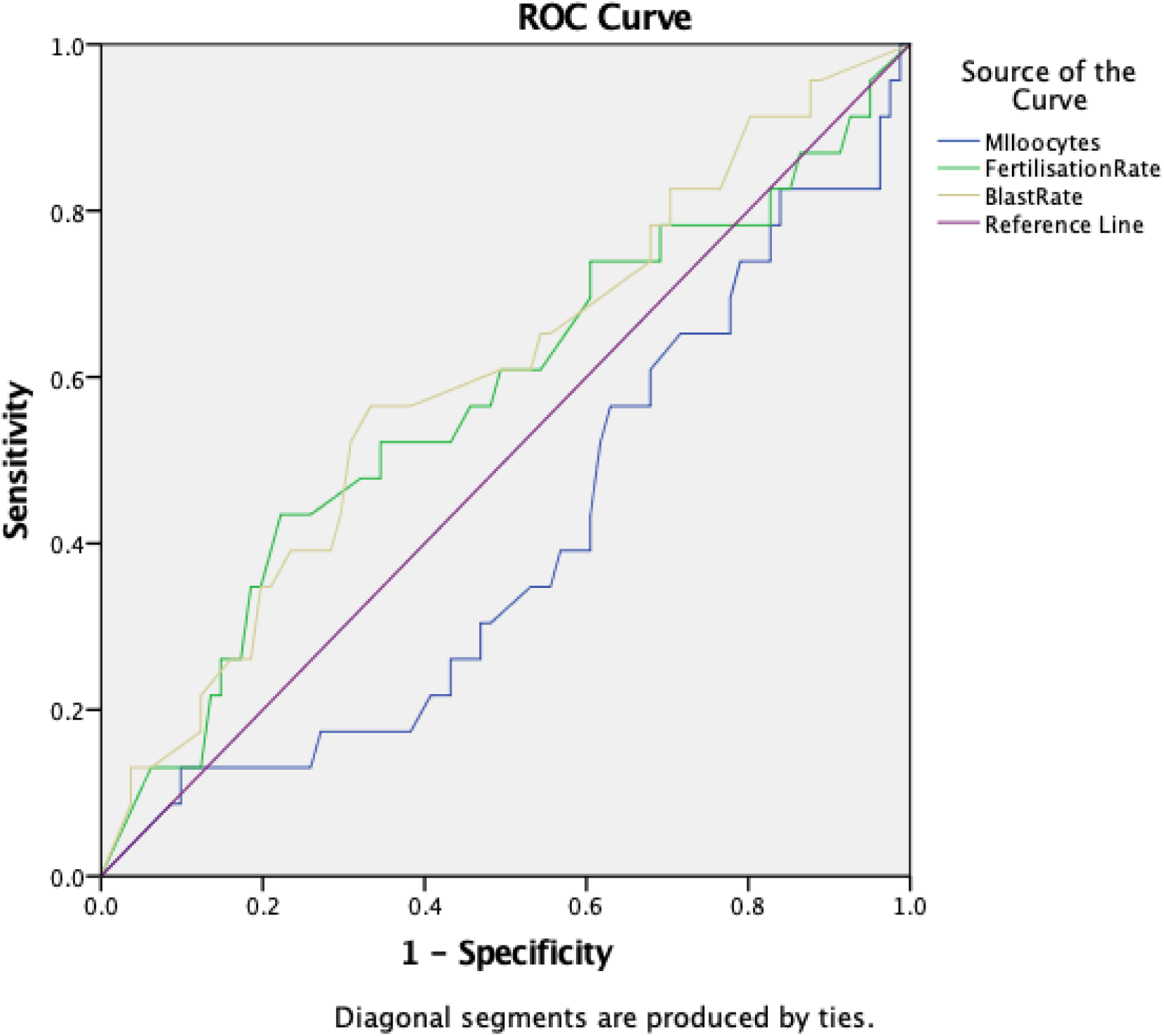
ROC curve showing predictive performance of LH levels, 12hrs post trigger, for percentage MII oocyte yield, fertilization rate and blastocyst conversion rate.

## DISCUSSION

We studied levels of LH, FSH, and progesterone in 104 women who underwent standard antagonist protocol IVF/ICSI cycles using GnRH agonist as a trigger. The levels of these hormones were checked at the time of the trigger and then 12 hours later, which correlates with peak serum titres. These levels were then correlated with mature oocyte yield, blastocyst yield, and fertilization rates. The primary end point was to determine LH levels 12 hours post trigger and correlate this with mature oocyte yield.

The results of this study showed that there is no statistically significant correlation between peak LH levels (12 hour post GnRH agonist trigger) and the development of mature oocytes. This was further demonstrated with ROC curve analysis showing limited ability of LH levels to predict mature oocyte yield. This contrasts data from earlier studies, such as by Shapiro et al. (15), where LH levels showed strong correlation with oocyte maturity. Our findings are in line with a retrospective cohort study by Dunne et al. (17), which showed no correlation between LH levels and mature oocyte yield (r=0.004). Furthermore, a recent study by Abbara et al. (18) shows similar findings in that peak LH levels showed little or no association to mature oocyte yield.

Shapiro et al. suggested a cut-off LH level of 12IU below which the yield of mature oocytes is suboptimal (15) and a systematic review of 37 studies by Ganer Herman et al. (19) suggested this cut-off be at 15IU. Furthermore, a more recent prospective cohort study by Blazquez et al. (20) disputes this cut-off of 15IU/L but rather suggests a new higher threshold of 22IU/L. Conversely, our data demonstrated that there is no LH level that we can set below which there is poor oocyte maturation. Of the data from the 103 patients that responded to COH, with a mature oocyte yield of 0-25%, the maximum LH level was 78.9IU/L and minimum was 41.9IU/L. In patients with mature oocyte yield of 25-50%, the maximum LH level was 79.8IU/L and minimum was 14.4IU/L. In patients with mature oocyte yield of 50-75%, the maximum LH level was 98.8IU/L and minimum was 14.2IU/L. In patients with mature oocyte yield of 75%-100%, the maximum LH level was 134.4IU/L and minimum was 6.8IU/L. The lowest mature oocyte yield was 4.5% and linked to a post-trigger LH level of 78.9IU/L. Furthermore, the highest mature oocyte yield was 100% and this was seen in 9 patients, associated with post-trigger LH levels as low as 6.8IU/L and as high as 81IU/L. Interestingly, 5 of these patients with 100% mature oocyte yield had LH levels below 16IU/L. As such, a specific minimum cut-off level for LH could not be determined below which oocyte maturation is unlikely to occur and the risk of empty follicle syndrome is increased.

hCG has been used for many decades as a surrogate for the physiological LH surge in order to induce oocyte maturation in IVF cycles. hCG is 6 to 7 times more biologically active than endogenous LH and also has a greater affinity for the LH receptor, resulting in prolonged and more potent luteotrophic effect in comparison (21). It is this quality that makes hCG a suitable and reliable trigger in IVF cycles. However, it is also this same property that results in ovarian hyperstimulation syndrome (22) and necessitates the need to find a suitable and safer alternative. GnRHa trigger has been introduced as this alternative as it reduces the risks of OHSS, but its reliability in initiating oocyte maturation is uncertain (23).

The main limitation of this study is the sample size included in the analysis and larger scale studies are needed to consolidate these findings. Additionally, studies have suggested that basal LH measurements rather than 12 hour post-trigger are a good predictor of outcomes when using GnRHa agonist trigger (24, 25). However, these studies have been largely retrospective and perhaps further prospective cohort studies are required. Perhaps there should be a focus on determining a more reliable predictor of mature oocyte yield. For example, the strongest positive correlation we found was with progesterone levels post-trigger. Similar findings have been shown in the study by Abbara et al. where progesterone rises strongly predicted the number of mature oocytes retrieved (18).

Reliably predicting sub-optimal oocyte yield in IVF cycles can offer a key benefit in avoiding undesirable outcomes as conditions can be optimised prior to egg collection and embryo transfer. In summary, our study did not show any specific level below which oocyte maturation, fertilisation, or blastocyst yields were significantly reduced. As such, routine measurement of LH level post GnRHa trigger cannot be recommended at this time to reliably predict IVF outcomes.

## Data Availability

All data produced in the present study are available upon reasonable request to the authors

## REFERENCES

1. Humaidan P, Ejdrup Bredkjær H, Bungum L, Bungum M, Grøndahl ML, Westergaard L, et al. GnRH agonist (buserelin) or hCG for ovulation induction in GnRH antagonist IVF/ICSI cycles: a prospective randomized study. Human Reproduction. 2005 May 1;20(5):1213–20.

2. Check JH, Nazari A, Barnea ER, Weiss W, Vetter BH. The efficacy of short-term gonadotrophin-releasing hormone agonists versus human chorionic gonadotrophin to enable oocyte release in gonadotrophin stimulated cycles. Human Reproduction. 1993 Apr;8(4):568– 71.

3. Alyasin A, Mehdinejadiani S, Ghasemi M. GnRH agonist trigger versus hCG trigger in GnRH antagonist in IVF/ICSI cycles: A review article. Int J Reprod Biomed. 2016 Sep;14(9):557– 66.

4. Delvigne A. Epidemiology and prevention of ovarian hyperstimulation syndrome (OHSS): a review. Human Reproduction Update. 2002 Nov 1;8(6):559–77.

5. Tummon I, Gavrilova-Jordan L, Allemand MC, Session D. Polycystic ovaries and ovarian hyperstimulation syndrome: a systematic review*. Acta Obstet Gynecol Scand. 2005 Jul;84(7):611–6.

6. Joham AE, Norman RJ, Stener-Victorin E, Legro RS, Franks S, Moran LJ, et al. Polycystic ovary syndrome. The Lancet Diabetes & Endocrinology. 2022 Sep;10(9):668–80.

7. Najdecki R, Michos G, Peitsidis N, Timotheou E, Chartomatsidou T, Kakanis S, et al. Agonist triggering in oocyte donation programs-Mini review. Front Endocrinol (Lausanne). 2022;13:838236.

8. Hernández ER, Gómez-Palomares JL, Ricciarelli E. No room for cancellation, coasting, or ovarian hyperstimulation syndrome in oocyte donation cycles. Fertility and Sterility. 2009 Apr;91(4):1358–61.

9. Kol S. Prediction of ovarian hyperstimulation syndrome: why predict if we can prevent! Human Reproduction. 2003 Jul 1;18(7):1557–8.

10. Kol S, Lewit N, ltskovitz-Eldor J. Ovarian hyperstimulation syndrome after using onadotrophin-releasing hormone analogue as a trigger of ovulation: causes and implications. Human Reproduction. 1996 Jun 1;11(6):1143–4.

11. Orvieto R. Can we eliminate severe ovarian hyperstimulation syndrome? Human Reproduction. 2005 Feb 1;20(2):320–2.

12. Itskovitz J, Boldes R, Levron J, Erlik Y, Kahana L, Brandes JM. Induction of preovulatory luteinizing hormone surge and prevention of ovarian hyperstimulation syndrome by gonadotropin-releasing hormone agonist. Fertil Steril. 1991 Aug;56(2):213–20.

13. Hoff JD, Quigley ME, Yen SSC. Hormonal Dynamics at Midcycle: A Reevaluation*. The Journal of Clinical Endocrinology & Metabolism. 1983 Oct;57(4):792–6.

14. Fauser BC, De Jong D, Olivennes F, Wramsby H, Tay C, Itskovitz-Eldor J, et al. Endocrine Profiles after Triggering of Final Oocyte Maturation with GnRH Agonist after Cotreatment with the GnRH Antagonist Ganirelix during Ovarian Hyperstimulation for *in Vitro* Fertilization. The Journal of Clinical Endocrinology & Metabolism. 2002 Feb;87(2):709–15.

15. Shapiro BS, Daneshmand ST, Restrepo H, Garner FC, Aguirre M, Hudson C. Efficacy of induced luteinizing hormone surge after “trigger” with gonadotropin-releasing hormone agonist. Fertility and Sterility. 2011 Feb;95(2):826–8.

16. Chen SL, Ye DS, Chen X, Yang XH, Zheng HY, Tang Y, et al. Circulating luteinizing hormone level after triggering oocyte maturation with GnRH agonist may predict oocyte yield in flexible GnRH antagonist protocol. Human Reproduction. 2012 May 1;27(5):1351–6.

17. Dunne C, Shan A, Nakhuda G. Measurement of Luteinizing Hormone Level After Gonadotropin-Releasing Hormone Agonist Trigger Is Not Useful for Predicting Oocyte Maturity. J Obstet Gynaecol Can. 2018 Dec;40(12):1618–22.

18. Abbara A, Hunjan T, Ho VNA, Clarke SA, Comninos AN, Izzi-Engbeaya C, et al. Endocrine Requirements for Oocyte Maturation Following hCG, GnRH Agonist, and Kisspeptin During IVF Treatment. Front Endocrinol (Lausanne). 2020;11:537205.

19. Ganer Herman H, Horowitz E, Mizrachi Y, Farhi J, Raziel A, Weissman A. Prediction, assessment, and management of suboptimal GnRH agonist trigger: a systematic review. J Assist Reprod Genet. 2022 Feb;39(2):291–303.

20. Blazquez A, Falcó N, Caño E, Rodriguez F, Vassena R, Miguel-Escalada I, et al. No association between LH levels and ovarian response in oocyte donors triggered with gonadotropin-releasing hormone agonist: A prospective study. Eur J Obstet GynecolReprod Biol. 2024 Mar;294:163–9.

21. Deepika K, Baiju P, Gautham P, Suvarna R, Arveen V, Kamini R. Repeat dose of gonadotropin-releasing hormone agonist trigger in polycystic ovarian syndrome undergoing In Vitro fertilization cycles provides a better cycle outcome - a proof-of-concept study. J Hum Reprod Sci. 2017;10(4):271.

22. Soares SR, Gomez R, Simon C, Garcia-Velasco JA, Pellicer A. Targeting the vascular endothelial growth factor system to prevent ovarian hyperstimulation syndrome. Human Reproduction Update. 2008 Apr 2;14(4):321–33.

23. Sun B, Ma Y, Li L, Hu L, Wang F, Zhang Y, et al. Factors Associated with Ovarian Hyperstimulation Syndrome (OHSS) Severity in Women With Polycystic Ovary Syndrome Undergoing IVF/ICSI. Front Endocrinol. 2021 Jan 19;11:615957.

24. Popovic-Todorovic B, Santos-Ribeiro S, Drakopoulos P, De Vos M, Racca A, Mackens S, et al. Predicting suboptimal oocyte yield following GnRH agonist trigger by measuring serum LH at the start of ovarian stimulation. Human Reproduction. 2019 Oct 2;34(10):2027– 35.

25. Lee WHY, Lin KT, Hsieh YC, Kao TC, Huang TC, Chao KH, et al. The value of LH maximum level in predicting optimal oocyte yield following GnRH agonist trigger. Front Endocrinol. 2023 Aug 7;14:1216584.

